# Analysis of 245,368 diverse individuals from the NIH All of Us Cohort identifies incomplete penetrance of the VEXAS-defining UBA1 p.M41L somatic mutation

**DOI:** 10.1101/2023.11.07.23298212

**Authors:** Robert W. Corty, Kevin Byram, Jason Springer, Peter C. Grayson, Alexander Bick

## Abstract

**Objective:** Somatic mutations in *UBA1* cause the recently described systemic auto-inflammatory syndrome, VEXAS. Study of this disease has largely been limited to highly symptomatic patients. We sought to determine the prevalence of VEXAS-associated somatic mutations and their disease penetrance in a diverse, unselected population.

**Methods:** We analyzed clinical-grade whole genome sequencing data from 245,368 individuals in the All of Us Research Program. We compared persons with canonical VEXAS-associated mutations to ten age, sex, and ancestry matched controls across the domains of diagnoses, medications, and laboratory values.

**Results:** 74 persons were identified with a VEXAS-defining somatic mutation at c.121A>C (p.Met41Leu) in *UBA1*. The variant allele fraction ranged from 4.5% to 33%. No other canonical VEXAS-associated mutations were identified. Of the 74 persons, 62 (84%) were women, 20 (27%) were African American, and 14 (19%) were American Admixed / Latino. There was no statistically significant association between case/control status and any diagnosis code, medication prescription, or laboratory value.

**Conclusion:** We report the largest cohort to date of persons with the VEXAS-associated p.Met41Leu mutation. This cohort differed substantially from reported cohorts of patients with clinical VEXAS, having a higher proportion of persons who were young, female, and of diverse ancestry. Variant allele fractions of p.Met41Leu mutations were lower than reported in clinical VEXAS and none of the patients had bioinformatically apparent VEXAS syndrome. The p.Met41Leu *UBA1* variant displayed incomplete penetrance for VEXAS. Further study is needed to determine the natural history of VEXAS-associated mutations in the pre-disease phase.

## Background

Somatic mutations in *UBA1* have recently been found to cause a systemic auto-inflammatory disease termed VEXAS syndrome, or simply VEXAS.^1^ VEXAS is named for five key characteristics of the disease: <u>v</u>acuoles are seen in the bone marrow, *UBA1* functions as an <u>E</u>1 enzyme in the ubiquitination pathway, *UBA1* is an <u>X</u>-linked gene, systemic <u>a</u>uto-inflammation drives most symptoms, and it is caused by <u>s</u>omatic mutation.

VEXAS was initially discovered in 2020 by a team studying patients referred for severe, recurrent auto-inflammation. Genetic analysis of these cohorts identified 25 men age 45 or older, each with one of three somatic mutations in *UBA1*: Met41Thr (60%), M41V (20%), and Met41Leu (20%).^1^

The initial description of the clinical syndrome included recurrent fever, skin involvement (neutrophilic dermatosis, leukocytoclastic vasculitis, and medium-vessel arteritis), pulmonary infiltrates, ear and nose chondritis, venous thromboembolism, macrocytic anemia, and vacuoles in hematologic precursor cells on bone marrow aspirates. Patients were noted to carry diagnoses of relapsing polychondritis, giant cell arteritis, Sweet syndrome, and myelodysplastic syndrome among others. Since that time, the spectrum of associated clinical manifestations has expanded to include: interstitial nephritis, cardiac involvement, and stroke in one cohort of twelve patients^2^, and genetically confirmed single patient reports of CIDP ^3^, AA amyloidosis ^4^, granulomatosis with polyangiitis ^5^, a lupus-like syndrome^6^, among others. The initial descriptions of VEXAS were confined to men in the fifth decade of life or later; however, the demographic spectrum has expanded to include a few cases in women with monosomy X^7–9^, euploid women^10^, and younger men^11^. Similarly, the range of genetic changes implicated in VEXAS syndrome has expanded to include intronic variants that affect splicing^8,12,13^, and missense mutations leading to Ser56Phe^12^, Ile894Ser^7^, Asn606Ile^7^, Ser621Cys^10^, and Gly477Ala.^14^

Recently, a study sought to identify the population prevalence and penetrance of VEXAS by identifying *UBA1* mutations in available exome sequencing data from a single health-system based cohort. Analysis of 163,096 mostly white patients identified 7 men with canonical (p.M41) VEXAS-associated somatic mutations, 3 men with other, likely pathogenic somatic mutations in *UBA1*, and two women with a somatic mutation in *UBA1* (one with a canonical mutation and one with non-canonical). Of the 12 patients, 6 had been diagnosed with VEXAS and all had features consistent with the disease.

Here we sought to evaluate the population prevalence and penetrance of VEXAS-associated somatic mutations in a racially, demographically, and geographically diverse cohort reflective of the full diversity of the United States.

## Methods

The *All of Us Research Program* is an NIH-funded program seeking to enroll one million or more persons in the United States of America who elect to share their electronic health record (EHR) data, donate biospecimens, and respond to surveys to create a nationwide prospective and retrospective population study, accelerate biomedical research, and improve health. To date, approximately 700,000 participants have enrolled. All of Us has performed clinical-grade, short-read whole genome sequencing in 245,368 individuals. Patients range in age from 18 to over 100 years. Approximately 75% have demographic features that are historically underrepresented in biomedical research related to race, ethnicity, age, sex, gender identity, sexual orientation, disability status, income, education, and geographic locality.

Somatic mutations were identified from short read sequences stored on the *All Of Us* data repository using the *Mutect2* tool in the GATK package and annotated with ANNOVAR.^15,16^ As described previously, variants were filtered to minimize technical artifacts based on: read depth >= 20, minimum allele depth >= 3, and at least one read from each direction corresponding to each allele.^17^ Somatic mutation at codon 41 in *UBA1* was prioritized based on an *a priori* strong association with VEXAS syndrome.

For each participant with somatic mutations at *UBA1* M41, controls were randomly selected at a 10:1 ratio from the sequenced cohort. Controls were matched on age, sex, and genetic ancestry and had no somatic mutations in *UBA1*.

The *All Of Us* program stores diagnosis codes harmonized into the SNOMED CT vocabulary. SNOMED CT codes were mapped to specific VEXAS manifestations as defined in the initial report and aggregated into related groups using R version 4.2.2. Medication prescriptions are stored in the *All Of Us* data repository using the RxNorm vocabulary. They were classified into drug classes based on custom, pragmatic rules using R version 4.2.2. Similarly, laboratory values are stored by *All Of Us* using the LOINC vocabulary. Lab values were filtered and transformed using custom, pragmatic rules in R. For each group of VEXAS-associated diagnosis codes, the Fisher exact test was used to compare the odds of having the code among cases against the odds among controls and test the null hypothesis that the odds were the same across genetically-defined groups. Analysis scripts are available at https://github.com/bicklab/UBA1_M41L_analysis.

## Results

We identified 74 participants in the NIH All Of Us Research Program with somatic mutation in *UBA1* at the M41 locus who we refer to as “cases”. Of the 74 cases, 62 (84%) reported their sex at birth to be female and genetic analysis found all of them to be euploid genetic females. The other 12 cases (16%) reported their sex at birth to be male and genetic analysis found all to be euploid genetic males. Cases ranged in age from 20 to 83 years at the time of sample collection and 21 to 87 years in 2023. Of the 74 cases, 39 (53%) reported white race, 17 (23%) reported black race, 2 (3%) reported Asian race, and the remainder declined to answer. Genetic analysis found that 38 (51%) were of primarily European ancestry, 20 (27%) were of primarily African or African American ancestry, 14 (19%) were of American Admixed / Latino ancestry, and one participant was of primarily East Asian and South Asian ancestry (Figure 1 and Table 1).

**Table 1:** characteristics of the 74 participants with somatic M41L mutation in context of matched controls, the large subset of the All Of Us cohort with whole genome sequencing data available, and the entire All Of Us cohort. Data is reported as mean and (standard deviation) or absolute number and (percent of cohort). Sex at birth, race, and ethnicity are self-reported. Genetic ancestry is computed based on genetic sequencing. Diagnosis codes and laboratory values are retrieved by the All Of Us program from participants’ Electronic Health Records data. Laboratory values underwent screening and filtering for interpretability and plausibility prior to inclusion in this study as detailed in *Methods*.

|  |  | Whole Genome Sequencing Cohort<br>(n = 245,368) | Female Controls<br>(n = 620) | Male Controls<br>(n = 120) | Female Cases<br>(n = 62) | Male Cases<br>(n = 12) |
| --- | --- | --- | --- | --- | --- | --- |
| Age at blood draw (years) [range] |  | 52 (17) [18, 114] | 49 (16) [20, 83] | 45 (15) [26, 69] | 49 (17) [20, 83] | 45 (15) [26, 69] |
| Genetic sex | XX | 150,261 | 620 (100%) | 0 | 62 (100%) | 0 |
|  | XY | 95,107 | 0 | 120 (100%) | 0 | 12 (100%) |
| Reported sex | Female | 145,550 (59%) | 606 (98%) | 0 | 62 (100%) | 0 |
|  | Male | 94,751 (39%) | 0 | 116 (97%) | 0 | 12 (100%) |
| Genetic ancestry | European | 133,566 (54%) | 309 (50%) | 70 (58%) | 31 (50%) | 7 (58%) |
|  | African / African American | 50,968 (23%) | 170 (28%) | 30 (25%) | 17 (27%) | 3 (25%) |
|  | American Admixed / Latino | 45,029 (18%) | 120 (20%) | 20 (17%) | 12 (19%) | 2 (17%) |
|  | East Asian | 5,706 (2%) | 10 (2%) | 0 | 1 (2%) | 0 |
|  | South Asian | 3,216 (1%) | 10 (2%) | 0 | 1 (2%) | 0 |
|  | Middle Eastern | 942 (0.4%) | 0 | 0 | 0 | 0 |
| race | White | 129,515 (53%) | 310 (50%) | 63 (53%) | 31 (50%) | 8 (67%) |
|  | Black or African American | 50,968 (21%) | 153 (25%) | 28 (23%) | 15 (24%) | 2 (17%) |
|  | Asian | 7,646 (3%) | 16 (3%) | 0 | 2 (3%) | 0 |
|  | Middle Eastern or North African | 1,389 (1%) | 1 (<1%) | 1 (<1%) | 0 | 0 |
| ethnicity | Not Hispanic or Latino | 188,637 (77%) | 481 (78%) | 92 (77%) | 47 (76%) | 10 (83%) |
|  | Hispanic or Latino | 47,367 (19%) | 122 (20%) | 21 (18%) | 13 (21%) | 2 (17%) |
| Diagnosis codes | Any diagnosis code | 179,380 (73%) | 469 (75%) | 89 (74%) | 49 (79%) | 10 (83%) |
|  | Length of EHR data (years) | 12 (8) | 11 (8) | 10 (8) | 12 (7) | 8 (3) |
|  | Number of unique codes | 42 (47) | 40 (43) | 34 (39) | 34 (37) | 18 (17) |
|  | 10%, 50%, and 90% quantiles of number of unique codes | [4, 26, 102] | [4, 26, 94] | [3, 19, 81] | [4, 21, 74] | [5, 12, 33] |
| Laboratory values | Any leukocyte concentration | 136,022 (55%) | 368 (59%) | 60 (50%) | 37 (60%) | 5 (42%) |
|  | Any lymphocyte % | 129,451 (53%) | 347 (56%) | 60 (50%) | 36 (60%) | 4 (33%) |
|  | Any hemoglobin concentration | 142,552 (58%) | 384 (62%) | 61 (51%) | 39 (65%) | 5 (42%) |
|  | Any MCV | 147,044 (60%) | 395 (64%) | 62 (52%) | 39 (65%) | 6 (50%) |
|  | Any ESR | 49,597 (20%) | 122 (20%) | 19 (16%) | 14 (23%) | 1 (8%) |
|  | Any CRP | 43,424 (18%) | 105 (17%) | 23 (19%) | 14 (22%) | 1 (8%) |
|  | Minimum leukocytes (cells per nL) | 5.8 (2.3) | 6.2 (2.9) | 6.1 (3.0) | 6.4 (2.7) | 6.1 (3.6) |
|  | Minimum lymphocytes (%) | 18 (11) | 18 (11) | 18 (11) | 19 (11) | 18 (15) |
|  | Minimum hemoglobin (mg/dL) | 11.5 (2.4) | 11.1 (2.2) | 12.7 (2.4) | 11.3 (2.0) | 13.0 (2.5) |
|  | Maximum MCV (fL) | 92 (6.3) | 91 (7) | 91 (8) | 92 (8) | 89 (3) |
|  | Maximum ESR (mm/hour) | 30 (30) | 29 (29) | 41 (41) | 30 (28) | 8 |
|  | Maximum CRP (mg/dL) | 38 (69) | 35 (65) | 55 (67) | 60 (109) | 18 |

**Figure 1a:**
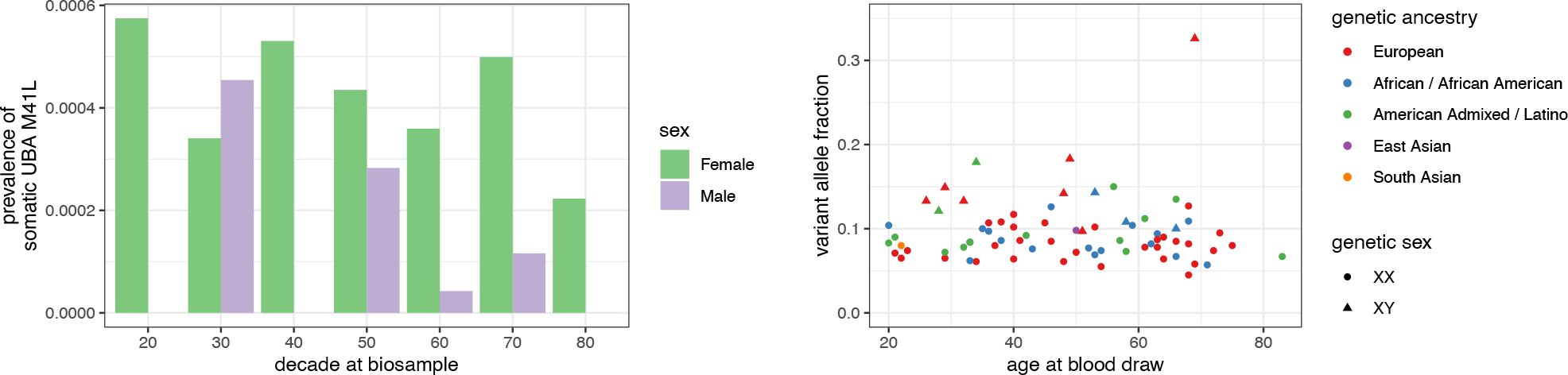
Somatic M41L prevalence is higher among women than men and, among women, is approximately constant from the 20’s to the 70’s. Among men, it is more common in the 30’s through 50’s. <u>Figure 1b</u>: Variant allele fraction of somatic M41L is similar across participants independent of age, genetic ancestry, and sex.

Of the 74 cases, all 74 had somatic DNA mutations at c.121A>C and therefore protein mutation p.Met41Leu. Manual review of the genetic locus revealed no repetitive sequences, as could induce systematic sequencing error. Their variant allele fraction ranged from 4.5% to 33% (Table 1).

There were no statistically significant differences in prevalence of VEXAS-associated diagnosis codes, nor in prescriptions for immunosuppressive medications, nor in VEXAS-associated laboratory abnormalities between cases and controls(Figure 2). However, cases did have a numerically higher rates of Behçet disease (2 of 74 vs. 0 of 740), Tietze’s disease (5 of 74 vs. 11 of 740), and systemic sclerosis (2 of 74 vs. 1 of 740).

**Figure 2a:**
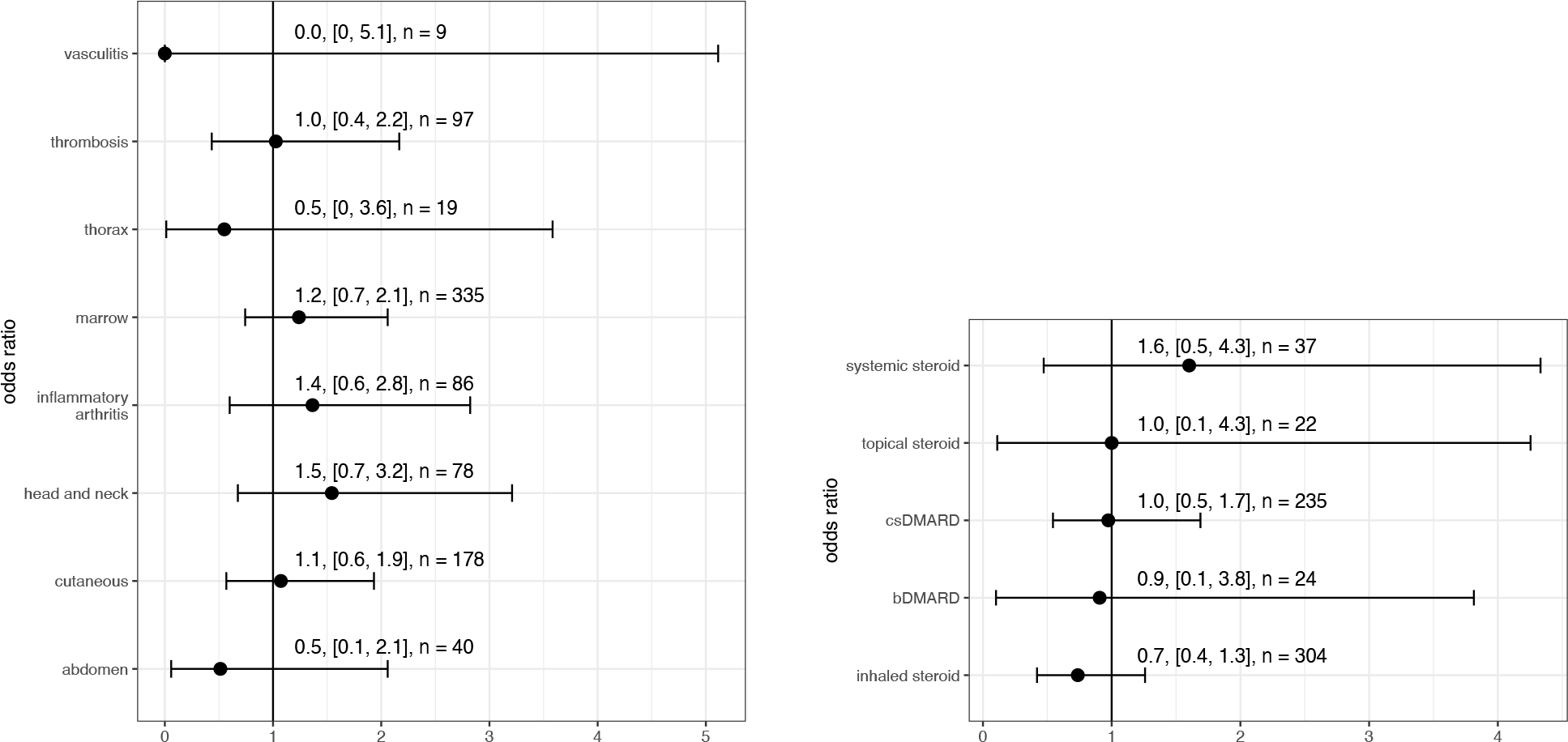
Participants with somatic p.Met41Leu and their matched controls have similar rates of VEXAS-associated diagnostic codes. All 9 occurrences of vasculitis were found among controls, so the point estimate for the odds ratio is 0, though the confidence interval includes 1. Details on diagnosis codes that define each category are available in the supplementary appendix. Not shown due to wide confidence intervals: pelvic manifestations, 0 among cases and 2 among controls yields odds ratio point estimate of 0 and confidence interval of [0, 53]. <u>Figure 2b</u>: Participants with somatic p.Met41Leu and their matched controls have similar rates of immunosuppressive medication prescription. DMARD: disease modifying anti-rheumatic drug. csDMARD: conventional synthetic DMARD. Not shown due to wide confidence intervals: calcineurin inhibitors (CNI) and targeted synthetic DMARDs (tsDMARD). CNI use was observed among 2 cases and 22 controls yielding an odds ratio of 0.91 with confidence interval [0.1, 3.8]. tsDMARD use was observed among 0 cases and 2 controls yielding an odds ratio of 0 with confidence interval [0, 53].

## Discussion

Analysis of the NIH All of Us cohort, identified 74 people with a single canonical VEXAS-associated mutation, representing the largest cohort of people with somatic mutation p.Met41Leu in the *UBA1* gene. The 62 women reported here represent the largest cohort of women with a VEXAS-associated mutation. This genetically-defined cohort did not have a significantly different rate of any specific diagnosis code. Nor did cases have different rates of medication prescription or VEXAS-associated lab abnormalities as compared to controls matched for age, sex, and genetic ancestry.

The clinical similarity between cases and matched controls is surprising in the context of the current understanding that the canonical VEXAS mutations are 100% penetrant in causing the VEXAS syndrome. This penetrance estimate, however, is based primarily on studies of patients experiencing severe, systemic, inflammatory disease and not on large, population-based studies. This selection bias was likely weaker in the published report of genetic screening for VEXAS mutations in a large health-system based cohort.^10^ However, that study involved mostly white and older patients who had at least some interaction with the U.S. healthcare system, which can introduce other subtle forms of selection bias. By leveraging the All Of Us Research Program, we have identified the largest and most diverse sample of individuals with a VEXAS-associated mutation that best represents the diversity of the United States, including many more women, young people, African American people, and Hispanic people as compared to existing cohorts.

The cohort reported here differs from other reported cohorts not only in demographic ways, but also on genetic parameters. In two large cohort studies of patients with VEXAS, p.Met41Thr was the most prevalent VEXAS defining mutation at codon 41 in *UBA1*, followed by p.Met41Val and p.Met41Leu. In this cohort, only variants in p.Met41Leu were detected. The p.Met41Leu variant has associated with less severe forms of VEXAS syndrome, thus it is reasonable that this variant may be over-represented a cohort that is generally much healthier than other cohorts of patients with UBA1 mutations. However, the absolute absence of other mutations despite the relatively large overall sample size suggests a countervailing form of selection bias may be as play in the All Of Us Research Program, where patients who are more severely ill may be under-represented. Additionally, the variant allele fraction (VAF) in this cohort, averaging 10%, is much lower than that of other cohorts, averaging 50%. We similarly interpret this lower VAF as consistent with the comparatively young and healthy cohort without clinical evidence of VEXAS syndrome, although even among patients with VEXAS syndrome, VAF is generally not correlated with disease severity.^13^

This study has multiple limitations. First, there may be diagnosis codes, prescriptions, and laboratory values associated with case or control patients that were not captured by the All Of Us Data Repository, most likely due to the fragmented nature of the U.S. medical system. Second, because the data collated in the All Of Us Data Repository is simply ingested from routine medical care, it is incomplete. Patients may have lab abnormalities that are not observed, diagnoses that are not documented either due to underdiagnosis or not having access to the relevant specialist or diagnostic test, and medications that were prescribed but never filled.

Although evidence is building as to the natural history of VEXAS syndrome, there have not been any studies on the natural history of VEXAS-associated somatic mutations in *UBA1*. Patients with such mutations could conceivably experience somatic reversion of the mutant clone and remain asymptomatic indefinitely. Alternatively, their mutant clone may persist and yet they may remain asymptomatic if the quantity of functional *UBA1* remains sufficient to perform needed ubiquitination functions. This eventuality seems more likely among women due to the X-linked nature of *UBA1*. And finally, the mutant clone could expand further and persons identified in this cohort could develop overt VEXAS syndrome. This cohort may enable the first pre-disease study of VEXAS syndrome, thereby advancing our understanding early steps in VEXAS pathogenesis that have, to date, been difficult to study. Until the natural history of VEXAS-associated somatic mutations can be clarified, it seems prudent to routinely monitor any people with such mutations. The genetic and clinical trajectory of these persons should be further characterized to inform strategies for risk stratification, monitoring and prompt treatment of VEXAS.

## Supporting information

SNOMED codes defining VEXAS manifestations

## Data Availability

All source data is available through the All of Us Researcher Workbench.

https://workbench.researchallofus.org/

## Acknowledgements

We appreciate scientific and editorial guidance from Drs. Leslie Crofford, MD, and C. Michael Stein, MBChB.

## Notes

### Competing Interest Statement

Unrelated to the current work, A.G.B. is a scientific co-founder and has equity in TenSixteen Bio. All other authors declare that they have no competing interests.

### Clinical Protocols

https://github.com/bicklab/UBA1_M41L_analysis

### Funding Statement

R.W.C. is supported by T32AR059039.
A.G.B. is supported by a Burroughs Wellcome Foundation Career Award for Medical Scientists and the NIH Director's Early Independence Award (DP5-OD029586).

### Author Declarations

IRB of All of Us Research Program, a program supported and overseen by the United States National Institutes of Health, gave ethical approval for this work

## References

1. Beck, D. B. et al. Somatic Mutations in UBA1 and Severe Adult-Onset Autoinflammatory Disease. N. Engl. J. Med. 383, 2628–2638 (2020).

2. van der Made, C. I. et al. Adult-onset autoinflammation caused by somatic mutations in UBA1: A Dutch case series of patients with VEXAS. J. Allergy Clin. Immunol. 149, 432-439.e4 (2022).

3. Bert-Marcaz, C. et al. Expanding the spectrum of VEXAS syndrome: association with acute-onset CIDP. J. Neurol. Neurosurg. Psychiatry 93, 797–798 (2022).

4. Euvrard, R. et al. VEXAS syndrome-related AA amyloidosis: a case report. Rheumatology 61, e15–e16 (2021).

5. Oka, H. et al. VEXAS syndrome with granulomatosis with polyangiitis manifestation: retained in remission using methotrexate and infliximab. Rheumatology (2023) doi:10.1093/rheumatology/kead536.

6. Valor-Méndez, L. et al. VEXAS syndrome mimicking lupus-like disease. Rheumatology 62, e271–e272 (2023).

7. Sakuma, M. et al. Novel causative variants of VEXAS in UBA1 detected through whole genome transcriptome sequencing in a large cohort of hematological malignancies. Leukemia 37, 1080–1091 (2023).

8. Templé, M. et al. Atypical splice-site mutations causing VEXAS syndrome. Rheumatology 60, e435–e437 (2021).

9. Barba, T. et al. VEXAS syndrome in a woman. Rheumatology 60, e402–e403 (2021).

10. Beck, D. B. et al. Estimated Prevalence and Clinical Manifestations of UBA1 Variants Associated With VEXAS Syndrome in a Clinical Population. JAMA 329, 318–324 (2023).

11. Sánchez-Hernández, B. E., Calderón-Espinoza, I. & Martín-Nares, E. Challenging the paradigm: a case of early-onset VEXAS syndrome. Rheumatology (2023) doi:10.1093/rheumatology/kead506.

12. Poulter, J. A. et al. Novel somatic mutations in UBA1 as a cause of VEXAS syndrome. Blood vol. 137 3676–3681 (2021).

13. Georgin-Lavialle, S. & Terrier, B. Further characterization of clinical and laboratory features in VEXAS syndrome: large-scale analysis of a multicentre case series of 116 French patients. British Journal of Dermatology (2022).

14. Stiburkova, B. et al. Novel Somatic UBA1 Variant in a Patient With VEXAS Syndrome. Arthritis Rheumatol 75, 1285–1290 (2023).

15. McKenna, A. et al. The Genome Analysis Toolkit: a MapReduce framework for analyzing next-generation DNA sequencing data. Genome Res. 20, 1297–1303 (2010).

16. Wang, K., Li, M. & Hakonarson, H. ANNOVAR: functional annotation of genetic variants from high-throughput sequencing data. Nucleic Acids Res. 38, e164 (2010).

17. Vlasschaert, C. et al. A practical approach to curate clonal hematopoiesis of indeterminate potential in human genetic data sets. Blood 141, 2214–2223 (2023).

